# Establishment of CORONET; COVID-19 Risk in Oncology Evaluation Tool to identify cancer patients at low versus high risk of severe complications of COVID-19 infection upon presentation to hospital

**DOI:** 10.1101/2020.11.30.20239095

**Authors:** R.J. Lee, C. Zhou, O. Wysocki, R. Shotton, A. Tivey, L. Lever, J. Woodcock, A. Angelakas, T. Aung, K. Banfill, M. Baxter, T. Bhogal, H. Boyce, E. Copson, E. Dickens, L. Eastlake, H. Frost, F. Gomes, D.M Graham, C. Hague, M. Harrison, L. Horsley, P. Huddar, Z. Hudson, S. Khan, U. T. Khan, A. Maynard, H. McKenzie, T. Robinson, M. Rowe, Anne Thomas, Lance Turtle, R. Sheehan, A. Stockdale, J. Weaver, S. Williams, C. Wilson, R. Hoskins, J. Stevenson, P. Fitzpatrick, C. Palmieri, D. Landers, T Cooksley, C. Dive, A. Freitas, A. C. Armstrong

**Author notes:** **Correspondence**: Dr Rebecca Lee, Department of Medical Oncology, The Christie NHS Foundation Trust, Wilmslow road, Manchester, M20 4BX. Equal contribution (R.J. Lee, C. Zhou, O. Wysocki). Equal contribution (C. Dive, A. Freitas, A. C. Armstrong).

## Abstract

**Background:** Cancer patients are at increased risk of severe COVID-19. As COVID-19 presentation and outcomes are heterogeneous in cancer patients, decision-making tools for hospital admission, severity prediction and increased monitoring for early intervention are critical.

**Objective:** To identify features of COVID-19 in cancer patients predicting severe disease and build a decision-support online tool; COVID-19 Risk in Oncology Evaluation Tool (CORONET)

**Method:** Data was obtained for consecutive patients with active cancer with laboratory confirmed COVID-19 presenting in 12 hospitals throughout the United Kingdom (UK). Univariable logistic regression was performed on pre-specified features to assess their association with admission (≥24 hours inpatient), oxygen requirement and death. Multivariable logistic regression and random forest models (RFM) were compared with patients randomly split into training and validation sets. Cost function determined cut-offs were defined for admission/death using RFM. Performance was assessed by sensitivity, specificity and Brier scores (BS). The CORONET model was then assessed in the entire cohort to build the online CORONET tool.

**Results:** Training and validation sets comprised 234 and 66 patients respectively with median age 69 (range 19-93), 54% males, 46% females, 71% vs 29% had solid and haematological cancers. The RFM, selected for further development, demonstrated superior performance over logistic regression with AUROC predicting admission (0.85 vs. 0.78) and death (0.76 vs. 0.72). C-reactive protein was the most important feature predicting COVID-19 severity. CORONET cut-offs for admission and mortality of 1.05 and 1.8 were established. In the training set, admission prediction sensitivity and specificity were 94.5% and 44.3% with BS 0.118; mortality sensitivity and specificity were 78.5% and 57.2% with BS 0.364. In the validation set, admission sensitivity and specificity were 90.7% and 42.9% with BS 0.148; mortality sensitivity and specificity were 92.3% and 45.8% with BS 0.442. In the entire cohort, the CORONET decision support tool recommended admission of 99% of patients requiring oxygen and of 99% of patients who died.

**Conclusions and Relevance:** CORONET, a decision support tool validated in hospitals throughout the UK showed promise in aiding decisions regarding admission and predicting COVID-19 severity in patients with cancer presenting to hospital. Future work will validate and refine the tool in further datasets.

## Introduction

The SARS-CoV-2 virus has infected over 30 million people to date, resulting in over a million deaths worldwide (1). A diverse spectrum of clinicopathological syndromes have been reported, ranging from asymptomatic cases to multi-organ failure and death (2). Standard medical care involves supportive therapies in those requiring hospital admission, with reduced mortality reported with dexamethasone in patients requiring oxygen or mechanical ventilation (3). Patients with milder symptoms have been safely managed as outpatients. Patients with cancer are at significantly increased risk of severe complications from COVID-19 including need for invasive ventilation and death (2,4). In two large case series and a meta-analysis of 18,650 patients, fatality rates of 10-30% have been observed in patients with cancer (5–7). Older age, male sex, Eastern Cooperative Oncology Group performance status (PS), smoking status, active cancer, haematological cancer and presence of other comorbidities such as hypertension have been shown to be significantly associated with mortality from COVID-19 (5–11).

Identifying oncology patients at risk of deterioration necessitating inpatient admission presents a unique challenge for healthcare professionals due to heterogeneity of clinical manifestations of COVID-19 and difficulty in distinguishing them from the complications of cancer and its therapy. In addition, to reduce burden on the health system and risk of nosocomial/hospital staff infection, it is important to admit only those patients who are likely to require additional supportive measures. A living review of risk prediction models has reported that current models are at high risk of bias and are poorly reported (12). More recently, the ISARIC 4C model has been developed using data from 57,824 patients in the United Kingdom (UK) to develop a score based on clinical/laboratory parameters (13). Although patients with a history of cancer were included in model development, it is unclear how well it performs in patients with active cancer.

We investigated clinical, haematological, and biochemical features in patients with active cancer presenting to hospital with COVID-19. Crucially, we wanted to create a pragmatic tool that can be readily applied in hospitals with parameters easily obtained through clinical history, examination and laboratory assessment. We developed a model, which aimed to predict whether cancer patients could be discharged safely without serious sequelae vs. severe disease requiring oxygen (O_2_) or death. Using this model we built an online tool; CORONET (COvid Risk Oncology Evaluation Tool) to support healthcare professionals in decisions regarding admission and to provide information as to the likely severity of illness. This is the first step of an iterative process whereby the tool will have ongoing refinement as more data and knowledge regarding COVID-19 and its treatment in cancer patients are obtained.

## Methods

### Study settings

Research Ethics Committee approval (reference 20/WA/0269) was granted to use a database of cancer patients presenting with COVID-19 (14) to establish the decision support tool and for follow-on data collection to be obtained prospectively, specifically for the CORONET project (see Supp methods).

### Data collection

Active cancer was defined as solid or haematological cancer diagnosed in the last 6 months or undergoing treatment for cancer or recurrent or metastatic cancer or haematological cancer not in complete remission for ≥6 months. Asymptomatic patients who were screened and found positive as part of routine testing for surgical procedures were not included due to lack of data. Patients had to have a laboratory confirmed SARS-CoV-2 infection (which for the majority was PCR-based).

### Selection of clinical, haematological, and biochemical features

Clinical, haematological and biochemical data were collected based on a pre-specified feature list including demographic/physiological features, oncology specific factors associated with poor cancer outcome such as performance status, literature review of features of COVID-19 severity and our previous work examining patients with cancer and COVID-19 longitudinally (14). Parameters were taken at presentation to hospital with symptoms of COVID-19, which was later laboratory confirmed or if already an inpatient, taken as close to/at the time of positive COVID-19 result (see Supp. methods for definitions of parameters).

### Patient outcomes

Admission (≥24 hours inpatient), O_2_ requirement and death were used as measures of infection severity. If outcomes were missing, the following assumptions were made: every patient that died due to COVID-19 also received O_2_ during hospital stay (4%, 11 pts missing O_2_), patients discharged within 24 hours did not receive O_2_ (<1%, 2 pts missing O_2_), patients that were inpatients for non-COVID reasons were also considered to be admitted (<1%, 2 pts missing O_2_), patients that died due to non-COVID reasons (deemed by clinician as asymptomatic of COVID-19 and dying of cancer) did not receive O_2_, patients admitted into ITU also received O_2_ (<1%, 2 pts missing O_2_).

### Study design

Transparent reporting of multivariable prediction models for individual prognosis or diagnosis (TRIPOD) guidelines have been used to report findings (15). Results from published data suggested that for parameters significantly associated with worse outcome in cancer patients with COVID-19, a sample size of 57 patients would provide a power of 90%, at a significance level of 0.05; for albumin minimum sample size = 28, CRP minimum sample size= 25, neutrophils minimum sample size = 57. In this study all statistical tests and modelling were carried out using R (ver 3.6.2) and Python (ver 3.7) (16,17).

### Model development

Model development workflow (Fig 1.) consisted of three stages: firstly an exploratory Pearson’s correlation analysis was performed to explore relationships between haematological and biochemical features. Univariable logistic regression analysis was carried out with each clinical/haematological/biochemical feature as an independent variable and each COVID-19 outcome as the dependent. This stage took place prior to model derivation, with the aim to screen for possible significant predictors for COVID-19 outcomes and explore the possible biology behind them. Features with p-values smaller than 0.1 were selected to go into the next analysis stage, except those with significant data missing (>50%). For patients with less than 2 missing features, missing values (haematological/biochemical only) were then imputed using multiple imputation.

**Figure 1.**
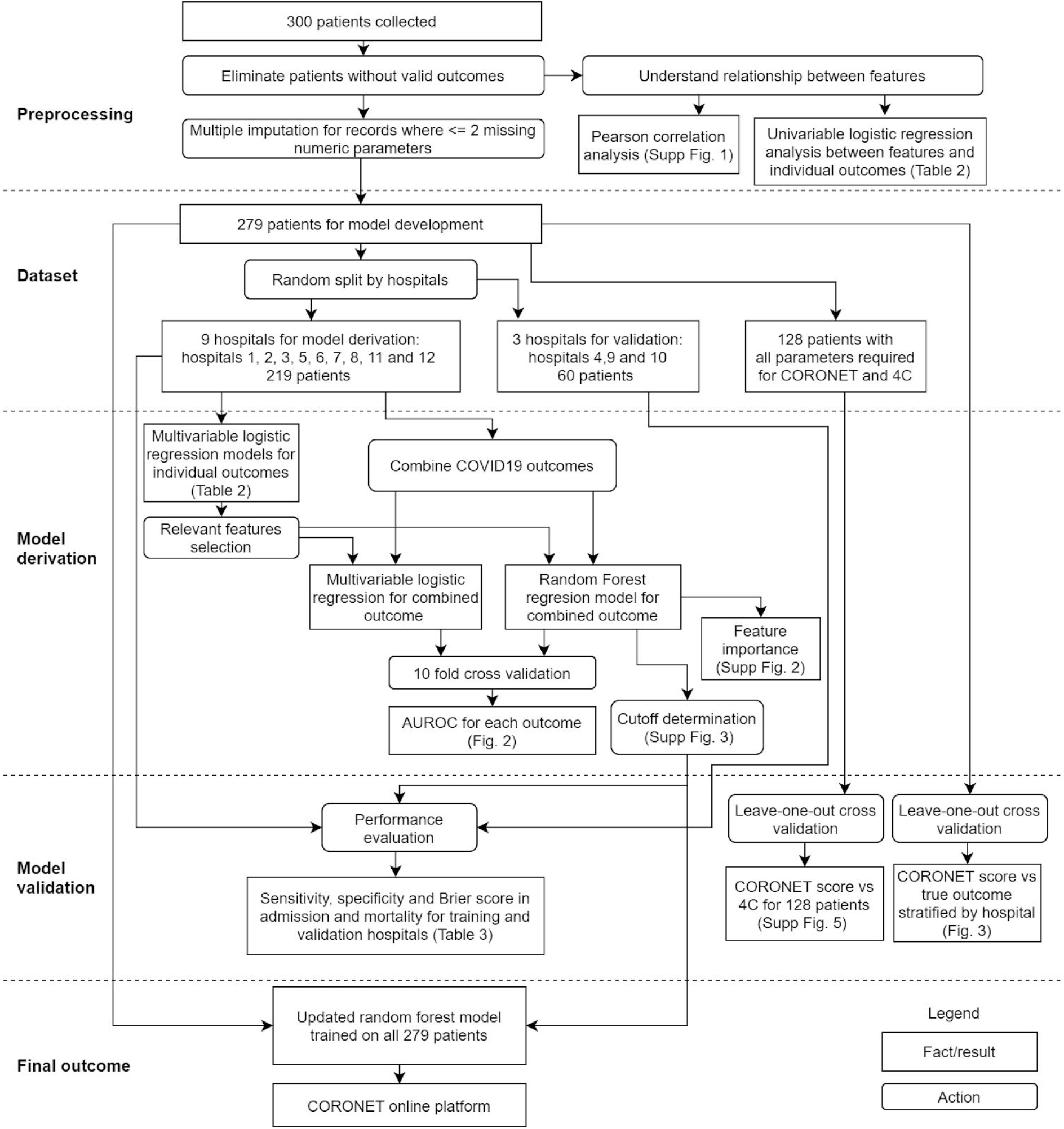
Modelling workflow. Model training and validation workflow. AUC= area under the curve, ROC= receiver operating curve, 4C= ISARIC 4C

Secondly, patients were randomly split into training (∼80%, 9 hospitals, 219 patients) and validation cohorts (∼20%, 3 hospitals, 60 patients) according to treating hospitals. Two different modelling approaches were applied: a multivariable logistic regression approach, selected due to model interpretability and its wide use in COVID-19 models currently available (13); plus a random forest approach (18), selected because of its superior theoretical accuracy as a machine learning method while still maintaining good interpretability compared to more sophisticated machine learning approaches.

The two models were developed in parallel and were compared in terms of both their statistical performance and clinical explainability. The final feature list for logistic regression was achieved using a backward stepwise method. Features with high correlation or with high biological relevance were manually inspected. Feature importance in the random forest model was evaluated using impurity (18). Model performance was evaluated by the area under receiver operating characteristic curve (AUROC) using 10-fold cross validation.

Finally, we identified optimal cut-offs to address key clinical questions namely necessity for admission due to likely severe COVID-19 and prediction of death. A cost function approach was adopted to address the imbalance of COVID-19 outcomes particularly surrounding admission where we observed a smaller number of patients being discharged (13%) vs. admitted (87% Table 1). The performance of the derived cut-offs were assessed by their sensitivity, specificity and Brier scores (19). In parallel we compared mortality prediction with the ISARIC 4C model and examined its utility in a cancer population (13).

**Table 1.**
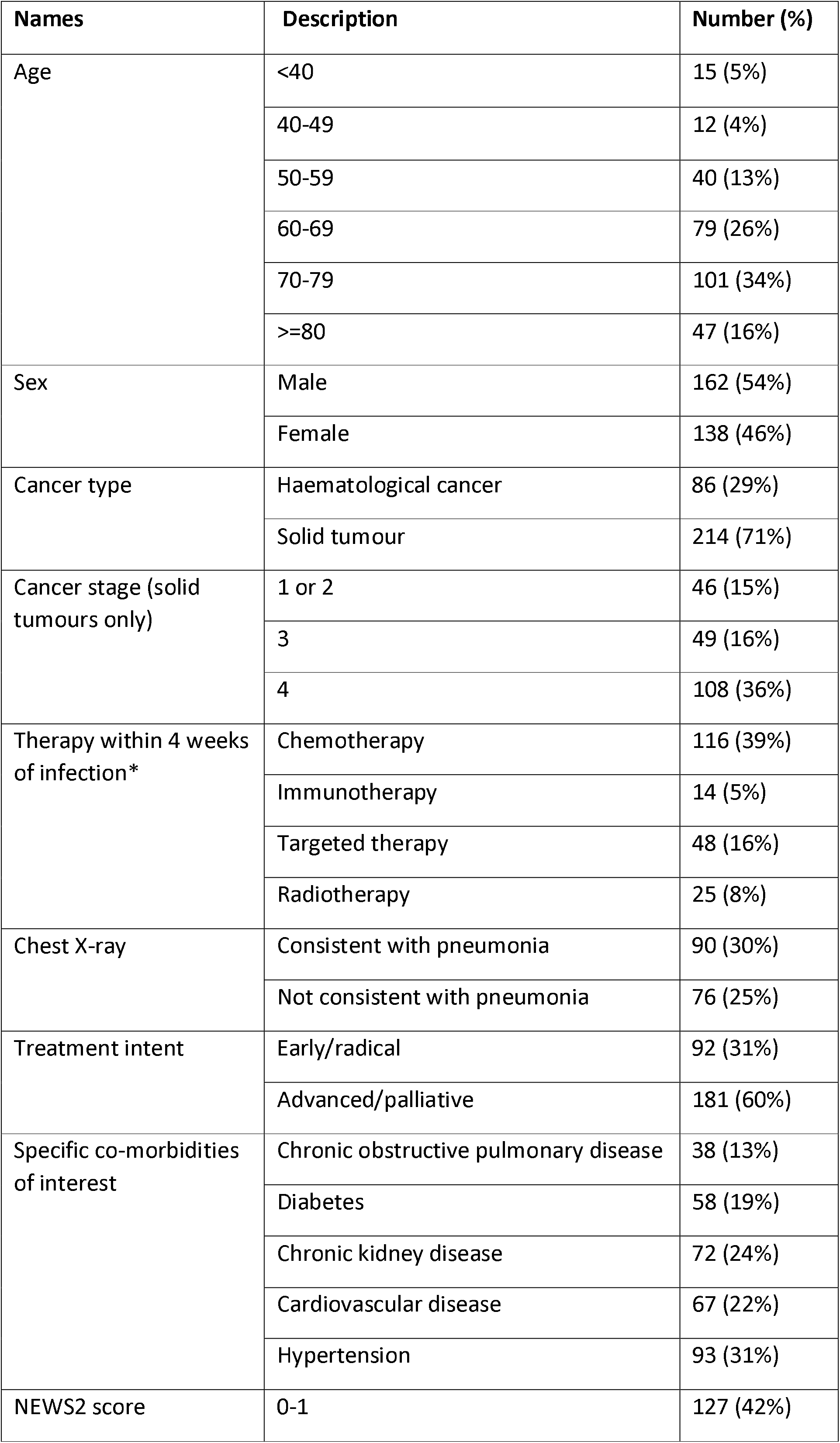
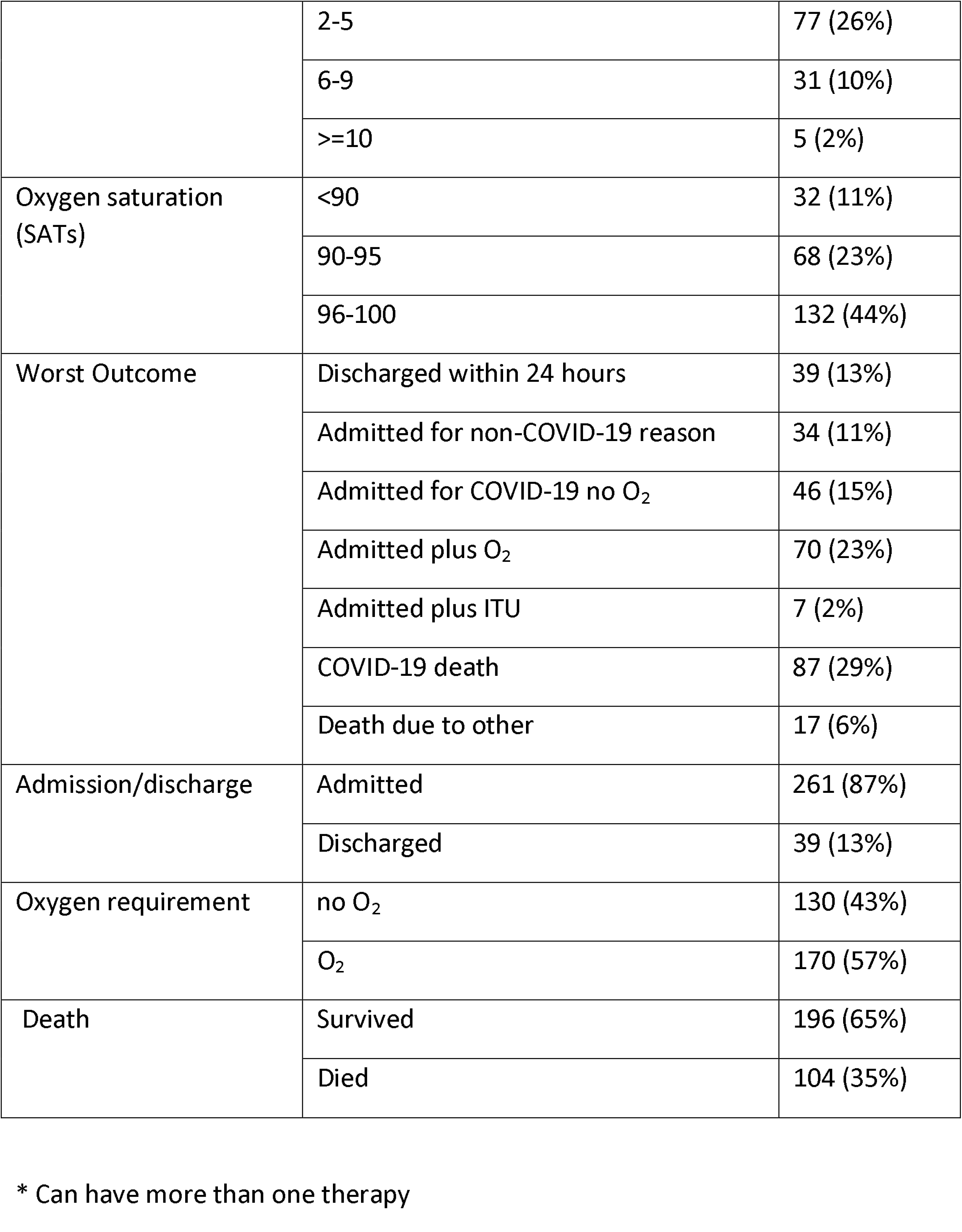
Summary of patients’ clinical characteristics.

| **Names** | **Description** | **Number (%)** |
| --- | --- | --- |
| Age | <40 | 15 (5%) |
|  | 40-49 | 12 (4%) |
|  | 50-59 | 40 (13%) |
|  | 60-69 | 79 (26%) |
|  | 70-79 | 101 (34%) |
|  | >=80 | 47 (16%) |
| Sex | Male | 162 (54%) |
|  | Female | 138 (46%) |
| Cancer type | Haematological cancer | 86 (29%) |
|  | Solid tumour | 214 (71%) |
| Cancer stage (solid tumours only) | 1 or 2 | 46 (15%) |
|  | 3 | 49 (16%) |
|  | 4 | 108 (36%) |
| Therapy within 4 weeks of infection* | Chemotherapy | 116 (39%) |
|  | Immunotherapy | 14 (5%) |
|  | Targeted therapy | 48 (16%) |
|  | Radiotherapy | 25 (8%) |
| Chest X-ray | Consistent with pneumonia | 90 (30%) |
|  | Not consistent with pneumonia | 76 (25%) |
| Treatment intent | Early/radical | 92 (31%) |
|  | Advanced/palliative | 181 (60%) |
| Specific co-morbidities of interest | Chronic obstructive pulmonary disease | 38 (13%) |
|  | Diabetes | 58 (19%) |
|  | Chronic kidney disease | 72 (24%) |
|  | Cardiovascular disease | 67 (22%) |
|  | Hypertension | 93 (31%) |
| NEWS2 score | 0-1 | 127 (42%) |
|  | 2-5 | 77 (26%) |
|  | 6-9 | 31 (10%) |
|  | >=10 | 5 (2%) |
| Oxygen saturation (SATs) | <90 | 32 (11%) |
|  | 90-95 | 68 (23%) |
|  | 96-100 | 132 (44%) |
| Worst Outcome | Discharged within 24 hours | 39 (13%) |
|  | Admitted for non-COVID-19 reason | 34 (11%) |
|  | Admitted for COVID-19 no O<sub>2</sub> | 46 (15%) |
|  | Admitted plus O<sub>2</sub> | 70 (23%) |
|  | Admitted plus ITU | 7 (2%) |
|  | COVID-19 death | 87 (29%) |
|  | Death due to other | 17 (6%) |
| Admission/discharge | Admitted | 261 (87%) |
|  | Discharged | 39 (13%) |
| Oxygen requirement | no O<sub>2</sub> | 130 (43%) |
|  | O<sub>2</sub> | 170 (57%) |
| Death | Survived | 196 (65%) |
|  | Died | 104 (35%) |

\* Can have more than one therapy

### Model validation and CORONET development

The derived model and suggested cut-offs were validated using the validation patient cohort, with sensitivity, specificity and Brier score to assess performance and calibration. Finally we updated and evaluated the model in the entire cohort of patients in order to provide the full data set to inform the CORONET online decision support tool.

## Results

### Clinical characteristics

Data collection was conducted between March-June 2020 from 12 participating hospitals, (mixture of local district general and tertiary centres, Supp. table 1.) throughout the United Kingdom (UK). Data for consecutive patients were obtained, with minimum 30 days follow up. Clinical features of all the patients are shown in Table 1. For the entire cohort, the median age was 69, range 19-93, with 54% males and 46% females and 71% having been diagnosed with a solid tumour whilst 29% had a haematological cancer. At the time of data cut-off, we grouped patients presenting with SARS-CoV-2 according to whether they had experienced 3 main outcomes associated with severity of COVID-19; admission to hospital (≥24 hours, 87% of patients), O_2_ (57% of patients) and death (29%). Very few patients (2%) were admitted to intensive care (ITU) therefore it was not used as an outcome measure for analysis.

### Association between variables and COVID-19 outcomes

Haematological and biochemical data are summarised in Supp. Table 2. Correlation between several biochemical and haematological measurements was low (median r=0.13, Supp. Fig. 1.), with maximum correlation observed between platelets and neutrophils (maximum r=0.49). We first performed univariable logistic regression analysis to assess the association between COVID-19 outcomes (admission/O_2_/death) and each clinical, biochemical and haematological variable (Table 2). A patient was significantly more likely to have severe COVID-19 outcomes (O_2_ and death) if they were older, male, had cardiovascular disease, high total number of comorbidities, haematological cancer advanced stage cancer (solid tumours examined only), poor performance status, high CRP, low albumin, high neutrophils, low oxygen saturation, high neutrophil/lymphocyte ratio (NLR), high LDH and high National Early Warning Score 2 (NEWS2 (20)) score (Table 2). Conversely, hospital admission was associated with a simpler set of variables: age, total number of comorbidities, CRP, albumin, SATs, LDH and NEWS2 score (Table 2). The discrepancy between significant features informing different outcomes was confirmed in multivariable logistic regression analysis (Table 2). For example, patients with haematological cancer had significantly higher risk of death (p=0.002) according to multivariate analysis, but it was not associated with increased hospital admission (p=0.327).

**Table 2.**
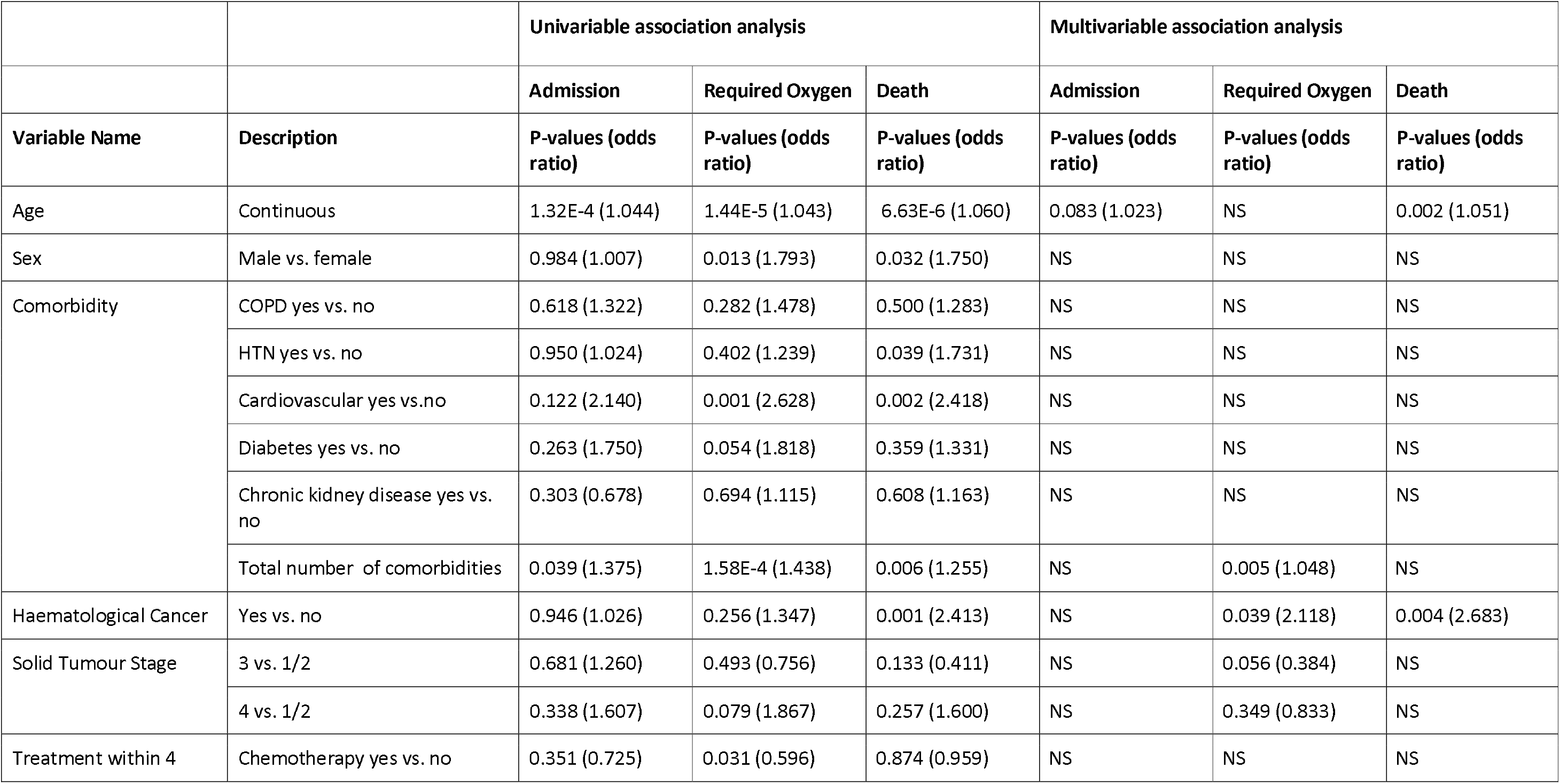
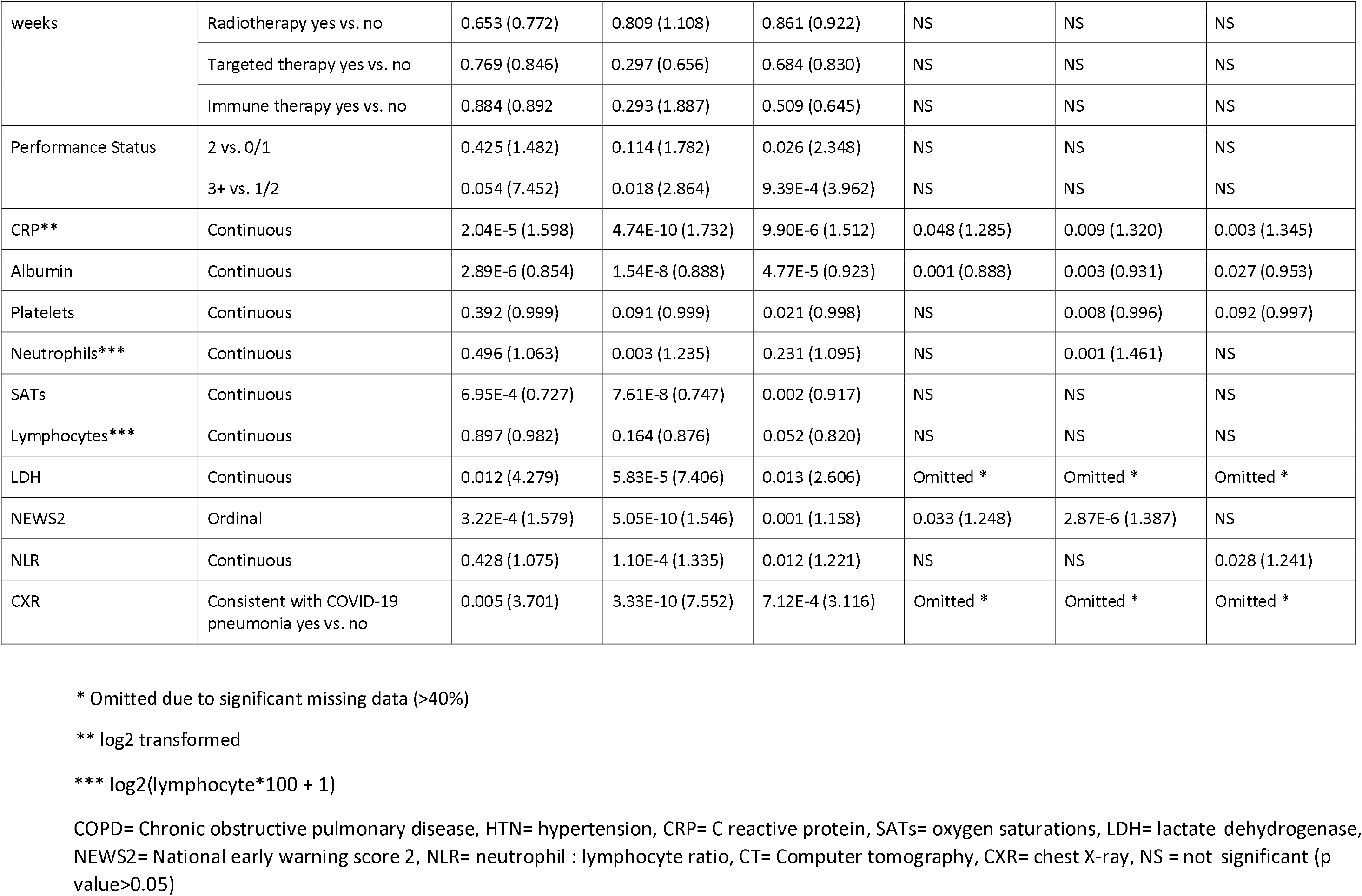
Association between COVID-19 outcomes and patient clinical/haematological/biochemical characteristics.

|  |  | Univariable association analysis |  |  | Multivariable association analysis |  |  |
| --- | --- | --- | --- | --- | --- | --- | --- |
|  |  | Admission | Required Oxygen | Death | Admission | Required Oxygen | Death |
| Variable Name | Description | P-values (odds ratio) | P-values (odds ratio) | P-values (odds ratio) | P-values (odds ratio) | P-values (odds ratio) | P-values (odds ratio) |
| Age | Continuous | 1.32E-4 (1.044) | 1.44E-5 (1.043) | 6.63E-6 (1.060) | 0.083 (1.023) | NS | 0.002 (1.051) |
| Sex | Male vs. female | 0.984 (1.007) | 0.013 (1.793) | 0.032 (1.750) | NS | NS | NS |
| Comorbidity | COPD yes vs. no | 0.618 (1.322) | 0.282 (1.478) | 0.500 (1.283) | NS | NS | NS |
|  | HTN yes vs. no | 0.950 (1.024) | 0.402 (1.239) | 0.039 (1.731) | NS | NS | NS |
|  | Cardiovascular yes vs.no | 0.122 (2.140) | 0.001 (2.628) | 0.002 (2.418) | NS | NS | NS |
|  | Diabetes yes vs. no | 0.263 (1.750) | 0.054 (1.818) | 0.359 (1.331) | NS | NS | NS |
|  | Chronic kidney disease yes vs. no | 0.303 (0.678) | 0.694 (1.115) | 0.608 (1.163) | NS | NS | NS |
|  | Total number of comorbidities | 0.039 (1.375) | 1.58E-4 (1.438) | 0.006 (1.255) | NS | 0.005 (1.048) | NS |
| Haematological Cancer | Yes vs. no | 0.946 (1.026) | 0.256 (1.347) | 0.001 (2.413) | NS | 0.039 (2.118) | 0.004 (2.683) |
| Solid Tumour Stage | 3 vs. 1/2 | 0.681 (1.260) | 0.493 (0.756) | 0.133 (0.411) | NS | 0.056 (0.384) | NS |
|  | 4 vs. 1/2 | 0.338 (1.607) | 0.079 (1.867) | 0.257 (1.600) | NS | 0.349 (0.833) | NS |
| Treatment within 4 | Chemotherapy yes vs. no | 0.351 (0.725) | 0.031 (0.596) | 0.874 (0.959) | NS | NS | NS |
| weeks | Radiotherapy yes vs. no | 0.653 (0.772) | 0.809 (1.108) | 0.861 (0.922) | NS | NS | NS |
|  | Targeted therapy yes vs. no | 0.769 (0.846) | 0.297 (0.656) | 0.684 (0.830) | NS | NS | NS |
|  | Immune therapy yes vs. no | 0.884 (0.892) | 0.293 (1.887) | 0.509 (0.645) | NS | NS | NS |
| Performance Status | 2 vs. 0/1 | 0.425 (1.482) | 0.114 (1.782) | 0.026 (2.348) | NS | NS | NS |
|  | 3+ vs. 1/2 | 0.054 (7.452) | 0.018 (2.864) | 9.39E-4 (3.962) | NS | NS | NS |
| CRP** | Continuous | 2.04E-5 (1.598) | 4.74E-10 (1.732) | 9.90E-6 (1.512) | 0.048 (1.285) | 0.009 (1.320) | 0.003 (1.345) |
| Albumin | Continuous | 2.89E-6 (0.854) | 1.54E-8 (0.888) | 4.77E-5 (0.923) | 0.001 (0.888) | 0.003 (0.931) | 0.027 (0.953) |
| Platelets | Continuous | 0.392 (0.999) | 0.091 (0.999) | 0.021 (0.998) | NS | 0.008 (0.996) | 0.092 (0.997) |
| Neutrophils*** | Continuous | 0.496 (1.063) | 0.003 (1.235) | 0.231 (1.095) | NS | 0.001 (1.461) | NS |
| SATs | Continuous | 6.95E-4 (0.727) | 7.61E-8 (0.747) | 0.002 (0.917) | NS | NS | NS |
| Lymphocytes*** | Continuous | 0.897 (0.982) | 0.164 (0.876) | 0.052 (0.820) | NS | NS | NS |
| LDH | Continuous | 0.012 (4.279) | 5.83E-5 (7.406) | 0.013 (2.606) | Omitted * | Omitted * | Omitted * |
| NEWS2 | Ordinal | 3.22E-4 (1.579) | 5.05E-10 (1.546) | 0.001 (1.158) | 0.033 (1.248) | 2.87E-6 (1.387) | NS |
| NLR | Continuous | 0.428 (1.075) | 1.10E-4 (1.335) | 0.012 (1.221) | NS | NS | 0.028 (1.241) |
| CXR | Consistent with COVID-19 pneumonia yes vs. no | 0.005 (3.701) | 3.33E-10 (7.552) | 7.12E-4 (3.116) | Omitted * | Omitted * | Omitted * |

\* Omitted due to significant missing data (>40%)
\*\* log2 transformed
\*\*\* $\log_2(\text{lymphocyte} \times 100 + 1)$
COPD= Chronic obstructive pulmonary disease, HTN= hypertension, CRP= C reactive protein, SATs= oxygen saturations, LDH= lactate dehydrogenase, NEWS2= National early warning score 2, NLR= neutrophil : lymphocyte ratio, CT= Computer tomography, CXR= chest X-ray, NS = not significant (p value>0.05)

### Modelling COVID-19 severity using combined outcomes

The discrepancy between factors significantly associated with different COVID-19 outcomes (admission vs. O_2_ and death) indicated that a decision on hospital admission needed to be improved by considering the risk of requiring O_2_ and death. Thus, a combined COVID-19 outcome was generated to represent severity, with multivariable logistic regression and random forest models subsequently developed using features significant in Table 2.

Performance of logistic regression and random forest models were evaluated by using them as a classifier to predict patient admission and death (Fig. 2). The random forest demonstrated superior performance with an AUC of ROC higher than that of logistic regression in both prediction of admission (0.85 vs. 0.78) and death (0.76 vs. 0.72) therefore was selected for further development. The importance of features involved in the random forest model are shown in Supp Fig 2, in which CRP was considered the most important feature to predict COVID-19 severity.

**Figure 2.**
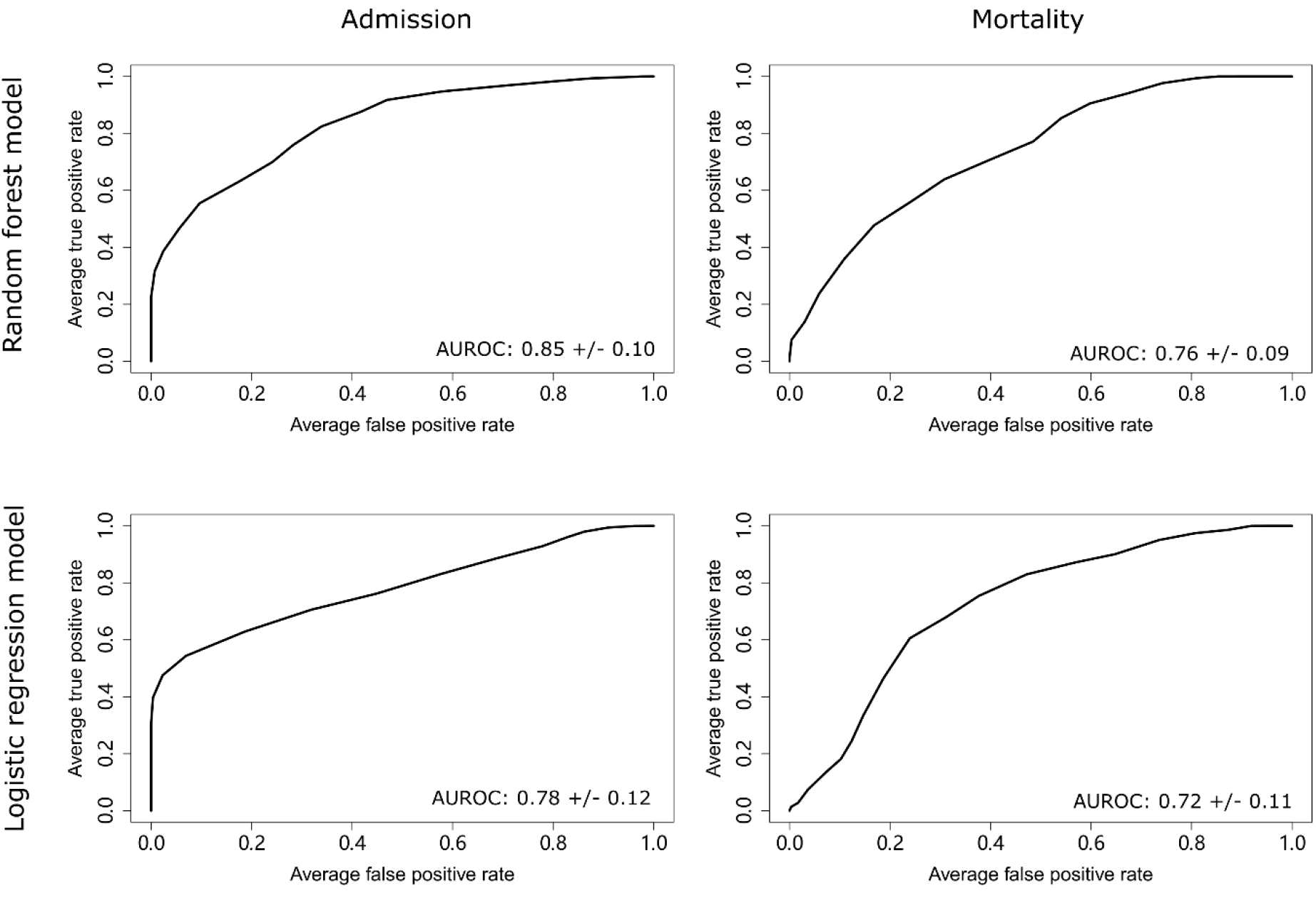
Performance of the random forest model and the logistic regression model in predicting patient admission and death. Area under receiving operator curve for logistic regression model and random forest model in predicting admission and mortality in patients presenting to hospital with COVID-19.

Cut-offs for predicting hospital admission and patient mortality were determined using a cost function approach to compensate for data imbalance (Supp Fig 3). The cut-off for admission was determined to be 1.05, while that for mortality being 1.8. At these thresholds, the model achieved a sensitivity of 78.5%, a specificity of 57.2%, and a Brier score of 0.364 in predicting patient mortality. Critically, for prediction of the need for admission it achieved a sensitivity of 94.5%, a specificity of 44.3%, and a Brier score of 0.118.

**Figure 3.**
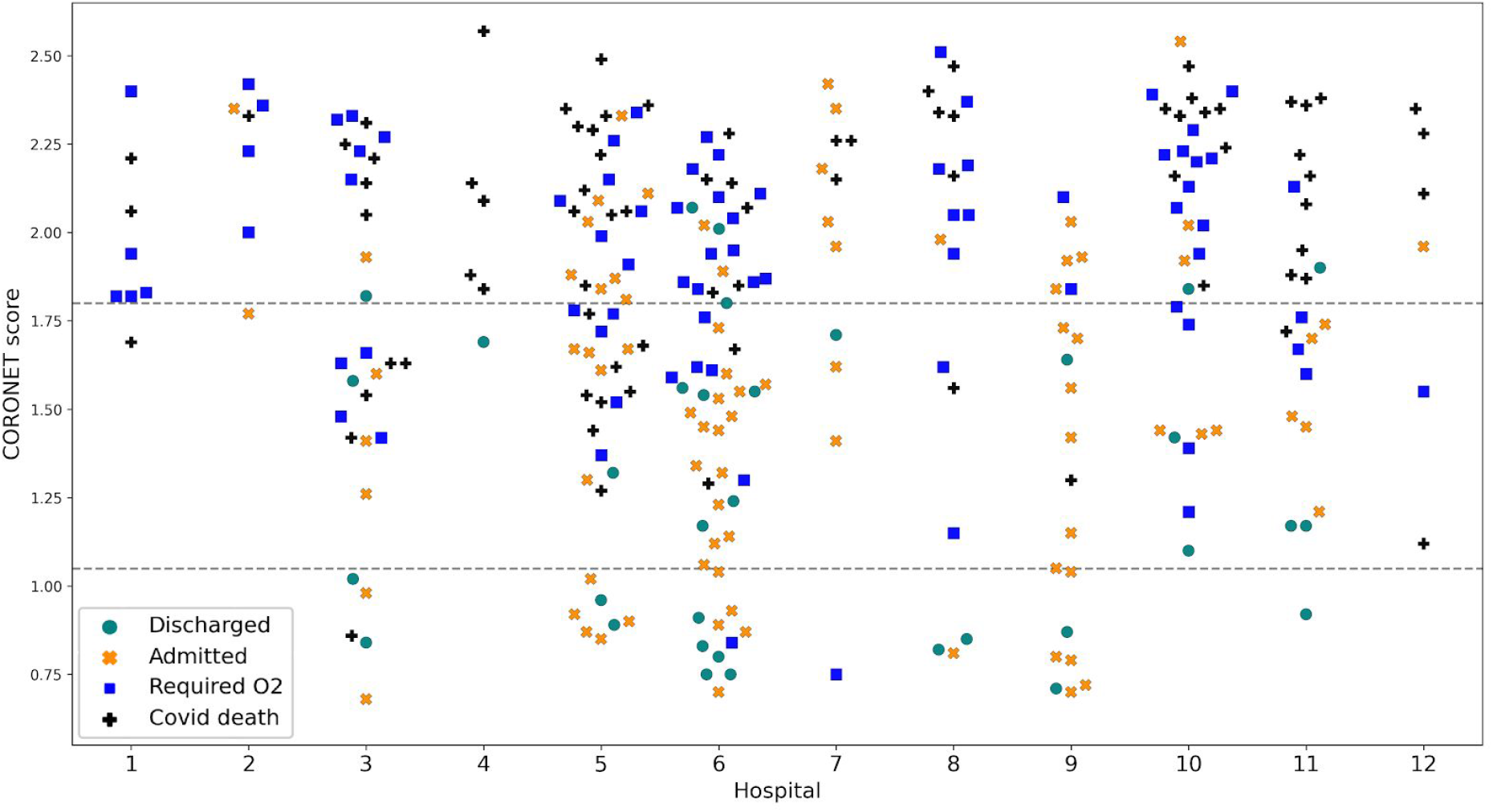
CORONET score and outcome in cancer patients presenting with COVID-19 to each hospital in the cohort. CORONET score calculated using Leave-One Out Cross Validation for 279 patients with cancer, stratified by hospital.

### Performance of the model in the validation cohort

Model performance was evaluated using the validation dataset (Supp. Table 3). The model achieved a consistent performance in admission with a sensitivity of 90.7%, a specificity of 42.9% and a Brier score of 0.148. For mortality, the model achieved a sensitivity of 92.30%, a specificity of 45.8% and a Brier score of 0.442.

### Comparison with ISARIC 4C score

Next, we identified a subset of 128 patients (named CORONET-4C), which had complete data available for parameters in both CORONET and ISARIC 4C mortality (4C (13)) models in order to compare them. In this subset, the AUROC for mortality for 4C was 0.81 and CORONET 0.74. Of note, 4C mortality scores in the CORONET-4C cohort were lower than the original 4C cohort, which was determined using predominantly non-cancer populations (Supp Fig. 4A). In addition, cancer patients were consistently more likely to die at lower values of the 4C score (Supp Fig. 4B). For example at a 4C score of ≤6, mortality was 4.5% in the original 4C validation cohort whereas in CORONET-4C it was 8.3% (Supp. Table 4). Critically, CORONET recommended admission of 100% of those patients who died and had 4C score ≤9, as well as 100% of patients with 4C ≥9. CORONET was trained on requirement for admission based on oxygen and death, which differed from the 4C model, which focused on risk of mortality. We therefore compared the 4C score rule in mortality threshold (≥9) and CORONET admission/severity thresholds (Supp. Figure 5). This revealed that the CORONET threshold safely admitted those patients who had required oxygen or died.

### Development of CORONET decision support tool

We applied the random forest model to patients in the entire cohort (n=279) and based on this developed the CORONET online platform. To demonstrate calibration of the updated CORONET model, its prediction of patient admission, O_2_ and mortality were plotted by hospital (Fig. 3). Critically, the CORONET score recommended admission for 99% of patients requiring O_2_ and 99% patients who died in the entire cohort, supporting its safety as a decision support tool.

## Discussion

Many studies have provided important data regarding risks of COVID-19 in the cancer population, which have helped to inform oncologists and patients in discussions regarding shielding, treatments and admissions with COVID-19 (5,6,8,10,11). We focused on developing a cancer-specific model of risk and decision support tool, which could aid the oncology and acute-care communities in discussions and decisions at the point of admission assessment of patients with symptoms of COVID-19.

In this cohort, the random forest model had the best performance with an AUROC of 0.85 for admission and 0.76 for death, which validated in different hospitals. Critically, in the entire cohort it recommended admission for 99% of patients who went on to require oxygen and 99% of the patients who died. In establishing our cut-offs, we prioritised achieving high sensitivity, which resulted in decreased specificity, but increased safety of the decision support tool. Modelling individual outcomes alone (e.g. death), may result in data over-fitting and does not reflect overall disease severity, therefore we chose to model a combined COVID-19 outcome. This may result in reduced accuracy on classification for a specific outcome, but improved generality to reflect COVID-19 severity, which was important for decision-making regarding hospital admission. We were also less stringent regarding those patients who were admitted but survived and did not receive oxygen, as these patients could potentially be managed at home. Patients may have been admitted due to oncological problems rather than COVID-19, therefore it is important to stress that the decision support tool is specific to COVID-19 rather than cancer-related decisions to admit.

Laboratory features such as CRP and clinical features such as age and haematological cancer have been shown to be independent risk factors by a number of groups (5,8,10,11). In other cohorts male sex and performance status have been identified as important independent negative prognostic factors; however these variables did not add to the predictive capability of our model (5,10). Intriguingly, haematological cancer and solid tumour stage were not as important as features such as CRP suggesting the COVID-19 induced inflammatory state is most critical in predicting severity. But, as patients with higher stage/haematological cancer are significantly more likely to develop severe COVID-19, these features are still useful for admission decisions. In addition, NEWS2 is commonly used within the UK to identify patients who are at risk of severe illness (20). Although NEWS2 has its own limitations and has been criticised especially in applicability to primary care (21), our validation of it as an important feature of severity in patients with cancer and COVID-19 suggests that it is helpful in the assessment of patients at least in the hospital setting.

We compared our model to the ISARIC 4C mortality risk score, created based on data from over 57,000 patients (13). Although a small cohort, it is important to note that our analysis of cancer patients using 4C showed that they were at higher risk of mortality with a lower 4C-score compared to the whole ISARIC population, which was mainly composed of patients without cancer. Thus, lower thresholds should be considered when using the 4C score in assessing patients with cancer. The 4C score had a better AUC for mortality compared to CORONET, which was likely due to our model training being focussed on admission of patients who were not only likely to die but also to require oxygen. In support of this, our model performed better in admission of patients requiring oxygen as a measure of COVID-19 severity. In addition, all those patients predicted by 4C to be at risk of mortality were admitted using our CORONET model, which is an important validation of its safety. Further comparisons in larger cohorts of patients are needed to better understand the benefits/limitations of each model in cancer patients.

There were several limitations in our model development. Firstly, the cohort is relatively small, however, according to the study design, it had sufficient power to detect significant differences in outcomes and validated across multiple hospitals. Through focusing on patients presenting to hospital, we selected a population biased towards more severe COVID-19. In addition, although we managed missing data through imputation and some assumptions (small numbers), we did not have sufficient data on features shown to be important in other cohorts such as ethnicity and LDH to incorporate into the analysis. These aspects will be addressed in future work validating the model further in international cohorts and prospective collection of additional parameters/data as more patients are admitted.

The majority of healthcare professionals have access to the internet and hospital results are increasingly accessed online. Consequently and in parallel, we have created a companion online decision tool (CORONET; COVID-19 risk in Oncology Evaluation Tool available at https://coronet.manchester.ac.uk/), which enables our model to be easily used. However, we recognise that for those working in resource poor settings this may provide a barrier to use and can provide further assistance if required. The tool is planned to provide prognostic information regarding the outcome of the patient in addition to assessment of how features of the individual define the outcome reported by the tool. In this way, we aim to support greater recognition of features that are associated with more severe outcomes for patients with cancer and COVID-19.

Critically, we view the creation and ongoing development of the decision support tool as an iterative process. This first version is a foundation upon which to improve as more data are obtained and more decision support features are created and validated in different hospitals. Using CORONET, healthcare professionals can be supported in their management of cancer patients with COVID-19. It aids discussions with patients and their families regarding likely prognosis, which is crucial to ensuring they are fully informed. It will support decisions regarding safe early discharge of patients, reducing hospital stay with beneficial impacts to emergency services, cost-savings and reducing risk of infecting staff/other patients. Furthermore, it will provide information, which can be used to identify those who might benefit from more intensive monitoring and to make early decisions regarding escalation to intensive care. In future, it may be used to identify patients at risk of severe COVID-19 who may have greatest benefit from interventions. Individualized management of COVID-19 presentations in cancer patients is crucial to providing sustainable emergency oncology care during the COVID-19 pandemic and beyond.

### Data availability

Code for the tool is available at Github (https://github.com/oskwys/CORONET). Raw data is available upon request to corresponding author, however may not include all details due to information governance regulations.

## Data Availability

https://coronet.manchester.ac.uk/

## Acknowledgements

Clare Griffin and Alison Backen for their support in the project. Digital ECMT team for insights into data analysis. Manchester and Liverpool ECMC.

