## Supplementary figures for "Establishment of CORONET; COVID-19 Risk in Oncology Evaluation Tool to identify cancer patients at low versus high risk of severe complications of COVID-19 infection upon presentation to hospital"

**Supplementary Figure 1. Correlation between haematological and biochemical features in cancer patients presenting to hospital with COVID-19**

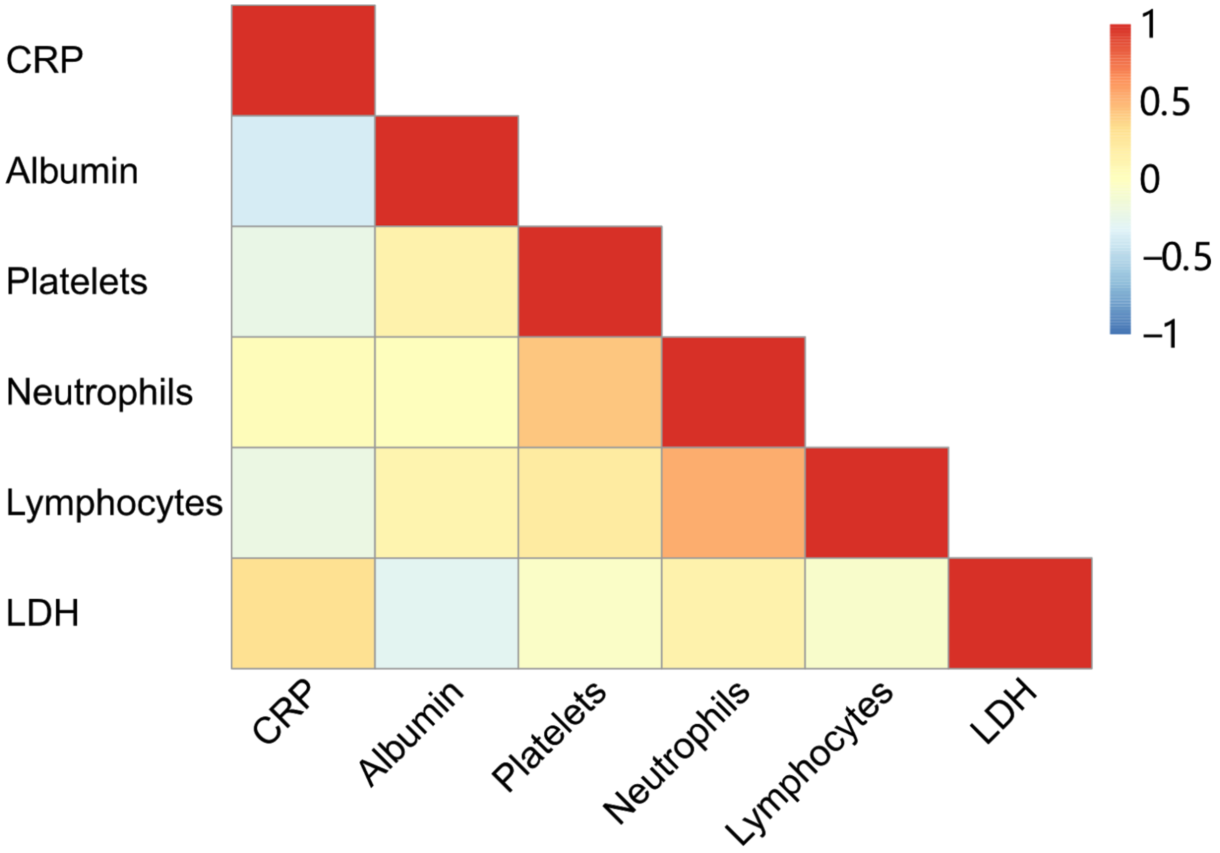


Pearson's correlation coefficients between haematological/biochemical features were calculated and plotted. Correlation levels were generally low with the median level at 0.13. The maximum correlation was observed between platelets and neutrophils at a level of 0.49. CRP=C-reactive protein, LDH= lactate dehydrogenase

**Supplementary Figure 2. Importance of features involved in the random forest model**

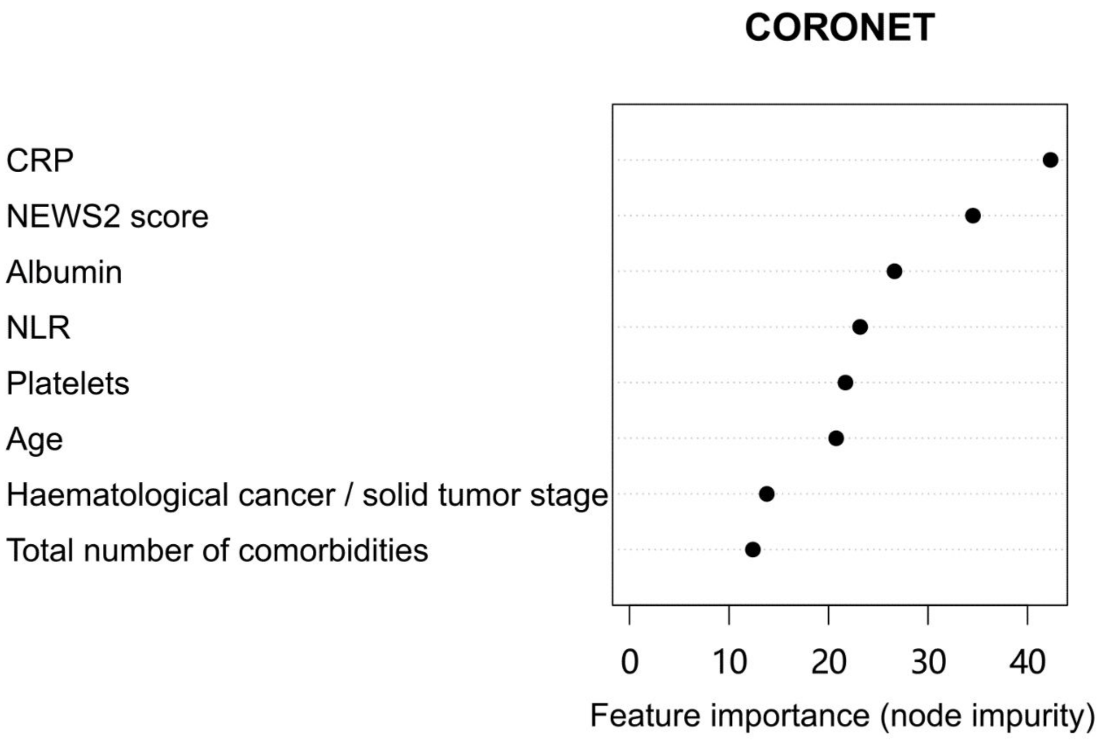


The importance of each feature in the random forest model were measured by node impurity. A feature is more important in predicting COVID-19 severity if it has high node impurity. CRP= C-reactive protein, NEWS2 score= National Early Warning Score 2, NLR= Neutrophil:lymphocyte ratio.

**Supplementary Figure 3. Cut-offs established in the random forest model to predict cancer patient admission and death**

A.

B.

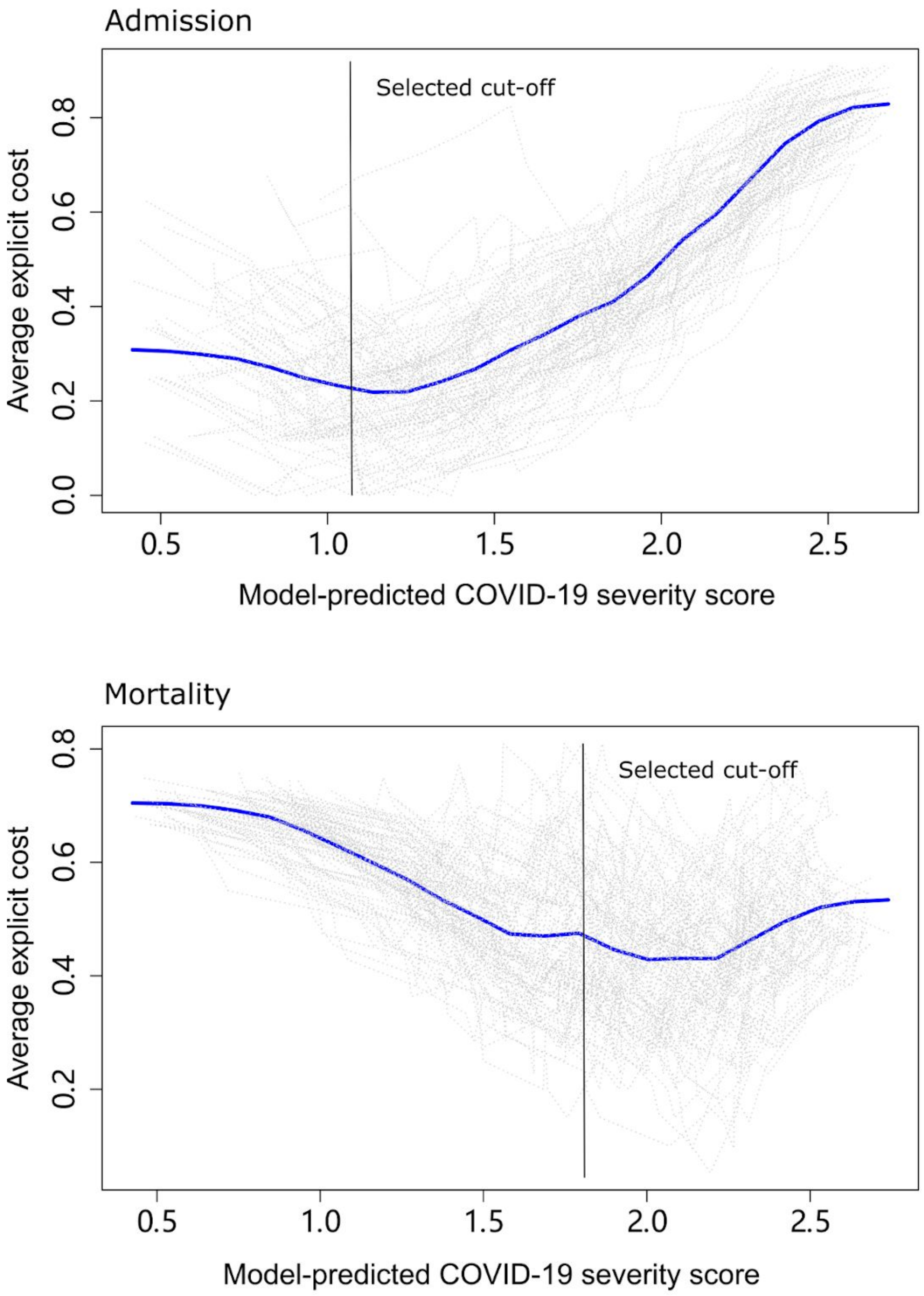


The cut-offs for predicting A. hospital admission and B. patient mortality were determined using a cost function approach to address data imbalance. Cut-offs were chosen to prioritise true positive prediction in order to ensure safety of prediction.

**Supplementary Figure 4. Analysis of 4C ISARIC model in cancer populations**

**A. Distribution of cancer patients vs. ISARIC cohort across 4C scores**

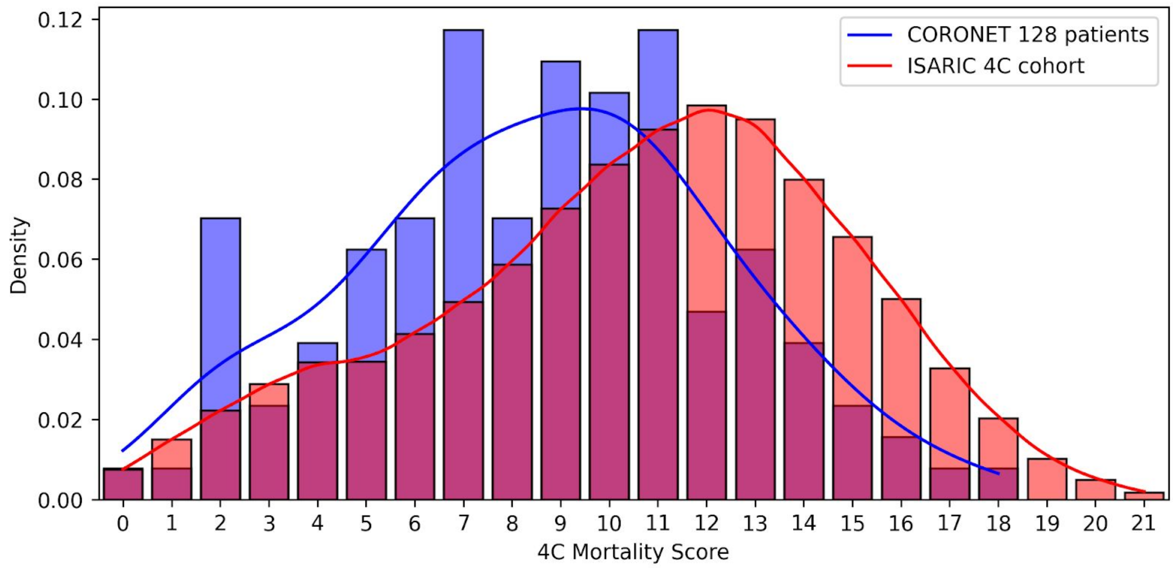


Distribution of patients across the ISARIC 4C mortality scores were examined in the CORONET-4C cohort (n=128) patients comprised of only cancer patients vs. the published ISARIC 4C cohort (n=22361), which comprised of a general admission patient population.

**B. Percentage mortality of cancer patients vs. ISARIC cohort across 4C scores.**

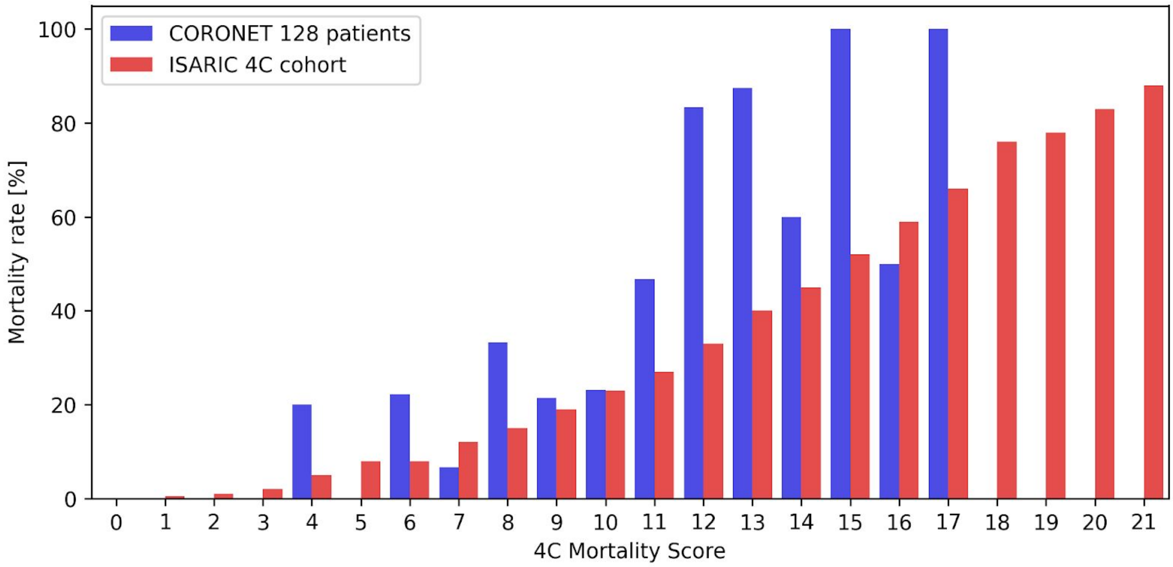


Mortality rate of patients across the ISARIC 4C mortality scores were examined in the CORONET-4C cohort (n=128) patients comprised of only cancer patients vs. the published ISARIC 4C cohort (n=22361), which comprised of a general admission patient population. Only deaths due to COVID-19 are considered.

**Supplementary Figure 5. Comparison of CORONET admission and severe condition thresholds with ISARIC 4C "Rule in mortality" threshold**

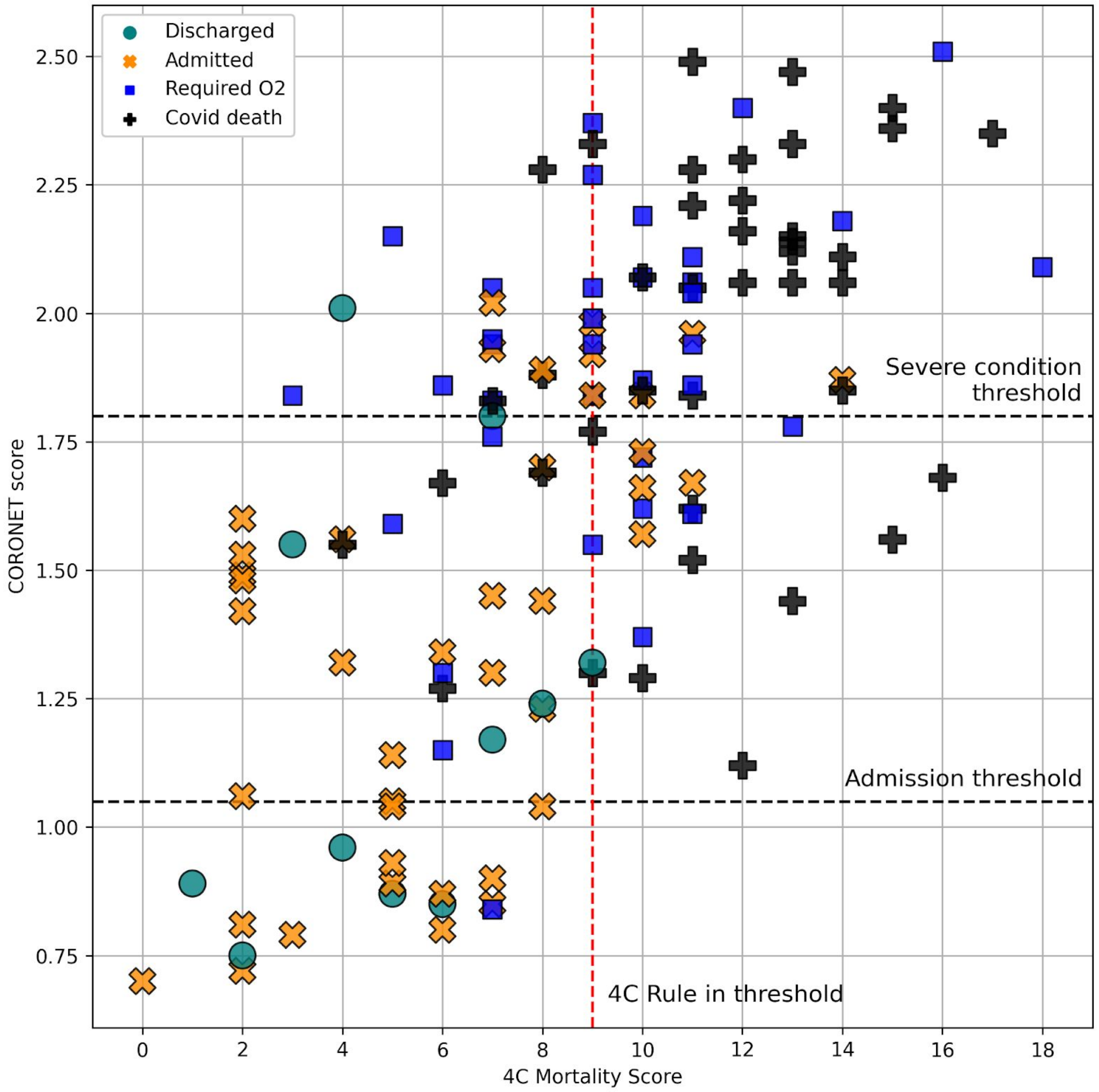


The ISARIC 4C rule in mortality threshold and CORONET admission and severe condition thresholds were compared in the CORONET-4C cancer population (n=128).
