## Supplementary tables for "Establishment of CORONET; COVID-19 Risk in Oncology Evaluation Tool to identify cancer patients at low versus high risk of severe complications of COVID-19 infection upon presentation to hospital"

**Supplementary Table 1. Number of patients admitted to each hospital**

| Hospital | # of patients |
| --- | --- |
| 1 | 8 |
| 2 | 7 |
| 3 | 29 |
| 4 | 6 |
| 5 | 53 |
| 6 | 62 |
| 7 | 12 |
| 8 | 19 |
| 9 | 21 |
| 10 | 33 |
| 11 | 23 |
| 12 | 6 |

**Supplementary Table 2. Summary of haematological and biochemical measurements**

| **Parameter** | **No of patients with measurements (%)** | **Mean (range)** |
| --- | --- | --- |
| C-reactive protein (CRP) | 259 (86%) | 99 (1-465) mg/L |
| Lymphocytes | 278 (93%) | 0.96 (0-18.9) x10^9^/L |
| Neutrophils | 280 (93%) | 5.5 (0-64.5) x10^9^/L |
| Neutrophil/lymphocyte ratio | 275 (92%) | 10.8 (0.03-222) |
| Platelets | 287 (96%) | 214 (8-773) x10^9^/L |
| Lactate dehydrogenase (LDH) | 95 (32%) | 402 (140-1379) IU/L |
| Albumin | 269 (90%) | 33.7 (10-51) g/L |

**Supplementary Table 3. Model validation performance with the proposed cut-off**

|  | **Training cohort** | | | **Validation cohort** | | |
| --- | --- | --- | --- | --- | --- | --- |
|  | Sensitivity | Specificity | Brier score | Sensitivity | Specificity | Brier score |
| Prediction for admission | 94.50% | 44.30% | 0.118 | 90.70% | 42.90% | 0.148 |
| Prediction for death | 78.50% | 57.20% | 0.364 | 92.30% | 45.80% | 0.442 |

**Supplementary Table 4. Comparison of ISARIC 4C Mortality score in original ISARIC 4C population and in the cancer population (CORONET-4C n=128 patients)**

| **4C Mortality score** | **No of patients (% of total 128)** | **TP** | **TN** | | **FP** | **FN** | **Sensitivity (%)** | **Specificity (%)** | **PPv (%)** | **nPv (%)** | **Mortality % in original ISARIC 4C publication (n=22361)** | **Mortality (%) in cancer population (n=128)** | **CORONET score (sd)** | **No of patients admitted by CORONET (%)** | **No of patients requiring O_2_ admitted by CORONET (%)** | **No of patients who died admitted by CORONET (%)** |
| --- | --- | --- | --- | --- | --- | --- | --- | --- | --- | --- | --- | --- | --- | --- | --- | --- |
| **Rule out mortality** | |  | |  | | | | | | | | | | | | |
| <=2 | 11 (8.6) | 40 | 11 | | 77 | 0 | 100.0 | 12.5 | 34.2 | 100.0 | 0.5 | 0.0 | 1.13 (0.37) | 6 (54.5) | 0 (-) | 0 (-) |
| <=3 | 14 (10.9) | 40 | 14 | | 74 | 0 | 100.0 | 15.9 | 35.1 | 100.0 | 1.2 | 0.0 | 1.19 (0.40) | 8 (57.1) | 1 (100) | 0 (-) |
| <=4 | 19 (14.8) | 39 | 18 | | 70 | 1 | 97.5 | 20.5 | 35.8 | 94.7 | 2.4 | 5.3 | 1.26 (0.41) | 12 (63.2) | 2 (100) | 1 (100) |
| <=5 | 27 (21.1) | 39 | 26 | | 62 | 1 | 97.5 | 29.5 | 38.6 | 96.3 | nd | 3.7 | 1.25 (0.41) | 16 (59.3) | 4 (100) | 1 (100) |
| <=6 | 36 (28.1) | 37 | 33 | | 55 | 3 | 92.5 | 37.5 | 40.2 | 91.7 | 4.5 | 8.3 | 1.24 (0.40) | 22 (61.1) | 9 (100) | 3 (100) |
| <=7 | 51 (39.8) | 36 | 47 | | 41 | 4 | 90.0 | 53.4 | 46.8 | 92.2 | nd | 7.8 | 1.34 (0.44) | 34 (66.7) | 15 (94) | 4 (100) |
| <=8 | 60 (46.9) | 33 | 53 | | 35 | 7 | 82.5 | 60.2 | 48.5 | 88.3 | 7.7 | 11.7 | 1.38 (0.44) | 42 (70) | 18 (95) | 7 (100) |
| <=9 | 74 (57.8) | 30 | 64 | | 24 | 10 | 75.0 | 72.7 | 55.6 | 86.5 | 10.0 | 13.5 | 1.48 (0.46) | 56 (75.7) | 27 (96) | 10 (100) |
| **Rule in mortality** | |  | |  | | | | | | | | | | | | |
| >=9 | 68 (53.1) | 33 | 53 | | 35 | 7 | 82.5 | 60.2 | 48.5 | 88.3 | 39.3 | 48.5 | 1.95 (0.33) | 68 (100) | 56 (100) | 33 (100) |
| >=10 | 54 (42.2) | 30 | 64 | | 24 | 10 | 75.0 | 72.7 | 55.6 | 86.5 | nd | 55.6 | 1.96 (0.32) | 54 (100) | 47 (100) | 30 (100) |
| >=11 | 41 (32) | 27 | 74 | | 14 | 13 | 67.5 | 84.1 | 65.9 | 85.1 | 44.5 | 65.9 | 2.03 (0.31) | 41 (100) | 38 (100) | 27 (100) |
| >=12 | 26 (20.3) | 20 | 82 | | 6 | 20 | 50.0 | 93.2 | 76.9 | 80.4 | nd | 76.9 | 2.07 (0.33) | 26 (100) | 25 (100) | 20 (100) |
| >=13 | 20 (15.6) | 15 | 83 | | 5 | 25 | 37.5 | 94.3 | 75.0 | 76.9 | 52.1 | 75.0 | 2.08 (0.30) | 20 (100) | 19 (100) | 15 (100) |
| >=14 | 12 (9.4) | 8 | 84 | | 4 | 32 | 20.0 | 95.5 | 66.7 | 72.4 | nd | 66.7 | 2.08 (0.30) | 12 (100) | 11 (100) | 8 (100) |
| >=15 | 7 (5.5) | 5 | 86 | | 2 | 35 | 12.5 | 97.7 | 71.4 | 71.1 | 61.5 | 71.4 | 2.14 (0.38) | 7 (100) | 7 (100) | 5 (100) |
| >=16 | 4 (3.1) | 2 | 86 | | 2 | 38 | 5.0 | 97.7 | 50.0 | 69.4 | nd | 50.0 | 2.16 (0.36) | 4 (100) | 4 (100) | 2 (100) |

nd – no data
