## Supplementary methods for "Establishment of CORONET; COVID-19 Risk in Oncology Evaluation Tool to identify cancer patients at low versus high risk of severe complications of COVID-19 infection upon presentation to hospital"

**Regulatory approvals**

Institutional approval was obtained following local information governance processes for case-note review at each site to establish a database to support wider clinical decision-making. In line with Health Research Authority (UK) guidance, database creation to support public health surveillance and clinical decisions is exempt from ethics committee review and anonymized data within a database can be used for research purposes if local governance approval is obtained (17). All patient data within the database was pseudo-anonymised with the key matching the study ID to patient identities kept at local NHS sites. Research Ethics Committee approval (reference 20/WA/0269) was granted to use data within the database for this study (which was anonymous to the researchers conducting this work), to establish the decision support tool.

**Hospitals included**

We aimed to include a range of hospitals managing cancer patients in different settings from local district general hospitals in which general acute physicians manage acute oncology admissions, to tertiary cancer centres where there are highly specialised oncology services.

**Parameter definitions**

*Total comorbidities* included chronic neurological/kidney/liver/rheumatological/cardio -vascular/respiratory/endocrine disease but did not include previous surgical procedures such as hip replacement.

*Cancer treatments* received within 4 weeks of presentation were categorised into chemotherapy, radiotherapy, immunotherapy and targeted therapy.

*Performance status* was classified according to Eastern Cooperative Oncology Group (ECOG) status
